# Prior infections and effectiveness of SARS-CoV-2 vaccine in test-negative study: A systematic review and meta-analysis

**DOI:** 10.1101/2022.11.03.22281925

**Authors:** Tim K. Tsang, Sheena G. Sullivan, Xiaotong Huang, Can Wang, Yifan Wang, Joshua Nealon, Bingyi Yang, Kylie E. C. Ainslie, Benjamin J. Cowling

**Affiliations:** WHO Collaborating Centre for Infectious Disease Epidemiology and Control, School of Public Health, Li Ka Shing Faculty of Medicine, The University of Hong Kong, Hong Kong Special Administrative Region, China; Laboratory of Data Discovery for Health Limited, Hong Kong Science and Technology Park, New Territories, Hong Kong Special Administrative Region, China; WHO Collaborating Centre for Reference and Research on Influenza, Royal Melbourne Hospital, and Doherty Department, University of Melbourne, at the Peter Doherty Institute for Infection and Immunity, Melbourne, Australia; Centre for Infectious Disease Control, National Institute for Public Health and Environment (RIVM), Bilthoven, the Netherlands

## Abstract

**Background:** Prior infection with SARS-CoV-2 can provide protection against infection and severe COVID-19. In settings with high pre-existing immunity, vaccine effectiveness (VE) should decrease with higher levels of immunity among unvaccinated individuals. Here, we conducted a systematic review and meta-analysis to understand the influence of prior infection on VE.

**Methods:** We included test-negative design (TND) studies that examined VE against infection or severe disease (hospitalization, ICU admission, or death) for primary vaccination series. To determine the impact of prior infections on VE estimates, we compared studies that excluded or included people with prior COVID-19 infection. We also compared VE estimates by the cumulative incidence of cases before the start of and incidence rates during each study in the study locations, as further measures of prior infections in the community.

**Findings:** We identified 67 studies that met inclusion criteria. Pooled VE among studies that included people with prior COVID-19 infection was lower against infection (pooled VE: 77%; 95% confidence interval (CI): 72%, 81%) and severe disease (pooled VE: 86%; 95% CI: 83%, 89%), compared with studies that excluded people with prior COVID-19 infection (pooled VE against infection: 87%; 95% CI: 85%, 89%; pooled VE against severe disease: 93%; 95% CI: 91%, 95%). There was a negative correlation between the cumulative incidence of cases before the start of the study and VE estimates against infection (spearman correlation (*ρ*) = −0.32; 95% CI: −0.45, −0.18) and severe disease (*ρ* = −0.49; 95% CI: −0.64, −0.30). There was also a negative correlation between the incidence rates of cases during the study period and VE estimates against infection (*ρ* = - 0.48; 95% CI: −0.59, −0.34) and severe disease (*ρ* = −0.42; 95% CI: −0.58, −0.23).

**Interpretation:** Based on a review of published VE estimates we found clear empirical evidence that higher levels of pre-existing immunity in a population were associated with lower VE estimates. Excluding previously infected individuals from VE studies may result in higher VE estimates with limited generalisability to the wider population. Prior infections should be treated as confounder and effect modificatory when the policies were targeted to whole population or stratified by infection history, respectively.

## INTRODUCTION

COVID-19 vaccines reduce the risk of infection and can also ameliorate disease severity when breakthrough infection occurs (1, 2). Ongoing evaluation of COVID-19 vaccine effectiveness (VE) has largely been measured through observational studies, particularly test-negative design (TND) studies, which share some similarities with case control studies (3). However, there has been substantial variation among reported VE estimates (4-7), which may be attributable to differences in study design, the vaccines used, disease incidence and population characteristics. Importantly, pre-existing population immunity as a result of infection could explain changes in COVID-19 VE over time and among populations (8, 9).

Infection with SARS-CoV-2 induces an immune response to protect against reinfection (10-14). However, reinfection could occur due to waning naturally-induced immunity (15, 16) or virus evolution (17, 18). Nevertheless, studies have shown that compared to persons with no prior infection, vaccination among people with prior infection enhances neutralising antibody activity as well as cell-mediated responses that can protect against (re)infection (19), suggesting prior infections may modify the protection from vaccinations. In settings where a large proportion of the population has prior exposure through infection, the unvaccinated will be more protected from infection than in a naïve population, thereby diluting the apparent effectiveness of vaccination. Under these two scenarios, prior infection modifies the effect of vaccination (Supplementary note 1).

Prior infection can also alter people’s decision to be vaccinated and present for care. For example, vaccination requirements vary for people with recent prior infection in Hong Kong (20). Moreover, individuals with recent infection may choose not to be vaccinated if they believe they have sufficient pre-existing immunity to prevent re-infection and ameliorate the severity of any re-infections that do occur (21). Additionally, these individuals may also choose not to present for care believing their COVID-like symptoms are due to another illness, leading to differential under-ascertainment of previously-infected COVID-19 cases in surveillance data. Other individual-level factors may also affect the decision to vaccinate and engage in infection-risk behaviors, such as perceived risk of severe disease post-infection (22-24). Acting in this way, prior infection may create a confounding bias along of the vaccination-COVID-19 association (Supplementary note 1).

Here, we aim to review systematically and meta-analyse published data to characterize the potential impact of pre-existing population immunity on VE estimates for completed primary vaccination series of COVID-19. We also conducted meta-regression to account for the influence of key design features such as vaccine types, circulating virus strains.

## METHODS

### Search strategy and selection criteria

This systematic review was conducted following the Preferred Reporting Items for Systematic Review and Meta-analysis (PRISMA) statement (25). A standardized search was done in PubMed, Embase and Web of Science, using the search term “(“test negative” OR “effectiveness”) AND (“vaccine”) AND (“COVID-19” OR “SARS-CoV-2”)”. The search was done on 11 July 2022, with no language restrictions. Additional relevant articles from the reference sections of identified articles were also reviewed. Two authors (XH and CW) independently screened the titles and full texts, and extracted data from the included studies, with disagreement resolved by consensus together with a third author (TKT). Studies identified from different databases were de-duplicated.

Studies that reported using a test-negative approach in which all cases and non-cases were tested were included (26, 27). We included published TND studies with participants recruited from the general population, and reported estimates of VE for completed primary vaccination series (two doses for most vaccines; one dose for Janssen) against at least one of the following endpoints: 1) positive test result, 2) symptomatic disease, 3) hospitalization, 4) ICU admission, 5) severe COVID, 6) death. We excluded articles if: 1) the study participants were recruited from a specific sub-population, such as healthcare professionals; 2) studies that only reported VE for booster doses; 3) studies that summarised or re-analysed already-published data; 4) studies that only reported pooled VE estimates for different vaccines; 5) the study was a preprint; or 6) the full text was not available.

Data were extracted from included studies using a standardised data collection form (Table S1) that collected information about: 1) study period; 2) region(s); 3) population; 4) the use of clinical criteria for enrolment; and 5) whether participants with prior SARS-CoV-2 infection were included. For each study, VE estimates with confidence intervals were extracted separately for each endpoint (e.g. infection, hospitalisation), vaccine and the circulating virus. In some studies, VEs specific to time intervals after vaccination were reported. Therefore, we extracted VE estimates for the first available time interval at least 14 days post-vaccination, because antibodies have been shown to peak by then in naïve persons (28). For studies that reported multiple estimates, such as by age group or type of vaccine, all subgroup-specific estimates were included, but the overall estimates were excluded.

### Meta-analysis

In all identified studies, VE was defined as 100%*(1-OR). The extracted VE estimates were meta-analysed to estimate pooled VE. VE estimates were transformed to the odds ratios scale, meta-analysed, then back-transformed to the VE scale for interpretation. The pooled odds ratio was estimated by random effects meta-analyses using the inverse variance method and restricted maximum likelihood estimator for heterogeneity (29-32). Heterogeneity was assessed using Cochran’s *Q* and the *I*^*2*^ statistic (33). We considered an *I*^*2*^ value more than 75% to be indicative of high heterogeneity (34). We also conducted a sensitivity analysis using fixed effects meta-analyses.

The main study feature of interest was if pooled VE against infection or severe disease varied depending on whether the studies included or excluded participants with prior infection. Severe disease was based on whether the estimate was limited to cases who required hospitalization, ICU admission and death. Otherwise the estimate was classified as VE against infection, which included estimates of VE against test positive or symptomatic infection (without hospitalisation).

Pooled estimates were additionally disaggregated by the probable circulating virus and vaccine administered. Most studies did not report variant-specific VE estimates but did report study periods and the general prevalence of variants during that period. Therefore, estimates were grouped according to the predominant circulating virus: 1) Omicron, 2) late-Delta, which was the period with co-circulation of Delta and Omicron, 3) Delta, 4) pre-Delta, which included ancestral strains and variants preceding Delta. Type of vaccine was grouped as follows: 1) mRNA vaccines, including vaccines produced by Moderna and Pfizer-BioNTech; 2) Adenovirus vector vaccines, including vaccine produced by AstraZeneca, Janssen and Gamaleya; and 3) Inactivated virus vaccines, including vaccine produced by Sinovac Biotech and Sinopharm.

### Meta-regression

To evaluate the impact of pre-existing immunity on VE estimates, we used a meta-regression approach. Three proxies of prior immunity were explored: 1) inclusion versus exclusion of participants with prior infection; 2) cumulative incidence of COVID-19 since December 2019 in each of the study countries/regions before the start of study; and 3) the incidence rate of COVID-19 in the country/region during the study period. For this, we downloaded population denominator data and daily COVID-19 case data from the World Health Organization website (35, 36). We first used correlation analysis, including Pearson (*r*) and Shearman (*ρ*) correlation coefficient, to determine the association between pre-existing immunity and VE estimates. Meta-regression models were adjusted for age group (age below or above 65 years), types of vaccines used, predominant circulating virus, and the use of clinical criteria for enrolment. A sensitivity analysis was conducted for additionally adjusting for location and duration of the study.

The fitted meta-regression model estimated the ratio of ORs (ROR) for each of the prior immunity proxies explored. On the OR scale, values closer to 0 indicated a more effective vaccine, while values closer to 1 indicated a less effective vaccine. This was counter to the VE scale where values closer to 0 indicated an ineffective vaccine. Therefore, using inclusion versus exclusion of participants with prior infection as an example, if ROR > 1, then the OR estimated from studies including participants with prior infection was higher than that from studies excluding participants with prior infection. On the VE scale, this translates to lower VE for studies that included participants with prior infection than studies that excluded these participants.

We plotted the expected change in VE estimate to visualize the impact of each prior immunity proxy based on the ROR obtained from meta-regression. To illustrate the change in VE scale, we showed the change in estimate based on the ROR assuming VE for the reference group of 80% against infection and 90% against severe disease. Statistical analyses were conducted using R version 4.0.5 (R Foundation for Statistical Computing, Vienna, Austria).

## RESULTS

We identified 6904 studies, among which 2929 were duplicates. Title and abstract screening of the remaining articles identified 480 for full text review, of which 67 met our inclusion criteria (4-7, 37-99) (Figure 1; Table S2). Studies were set in 17 countries/regions. Most were from the United States (29) and United Kingdom (10). Fifty-one studies provided 173 VE estimates against infection (Figure S1-2), and 41 studies provided 93 estimates against severe disease (Figure S3-4). Among all 67 studies, 45 included and 24 studies excluded participants with prior COVID-19 infection (including two studies which provided VE estimates including and excluding participants with COVID-19 infection). A summary of study characteristics and the corresponding number of estimates, including handling of participants with prior infections, enrolment criteria, vaccine types and circulating virus are provided in Table S3-S5.

**Figure 1.**
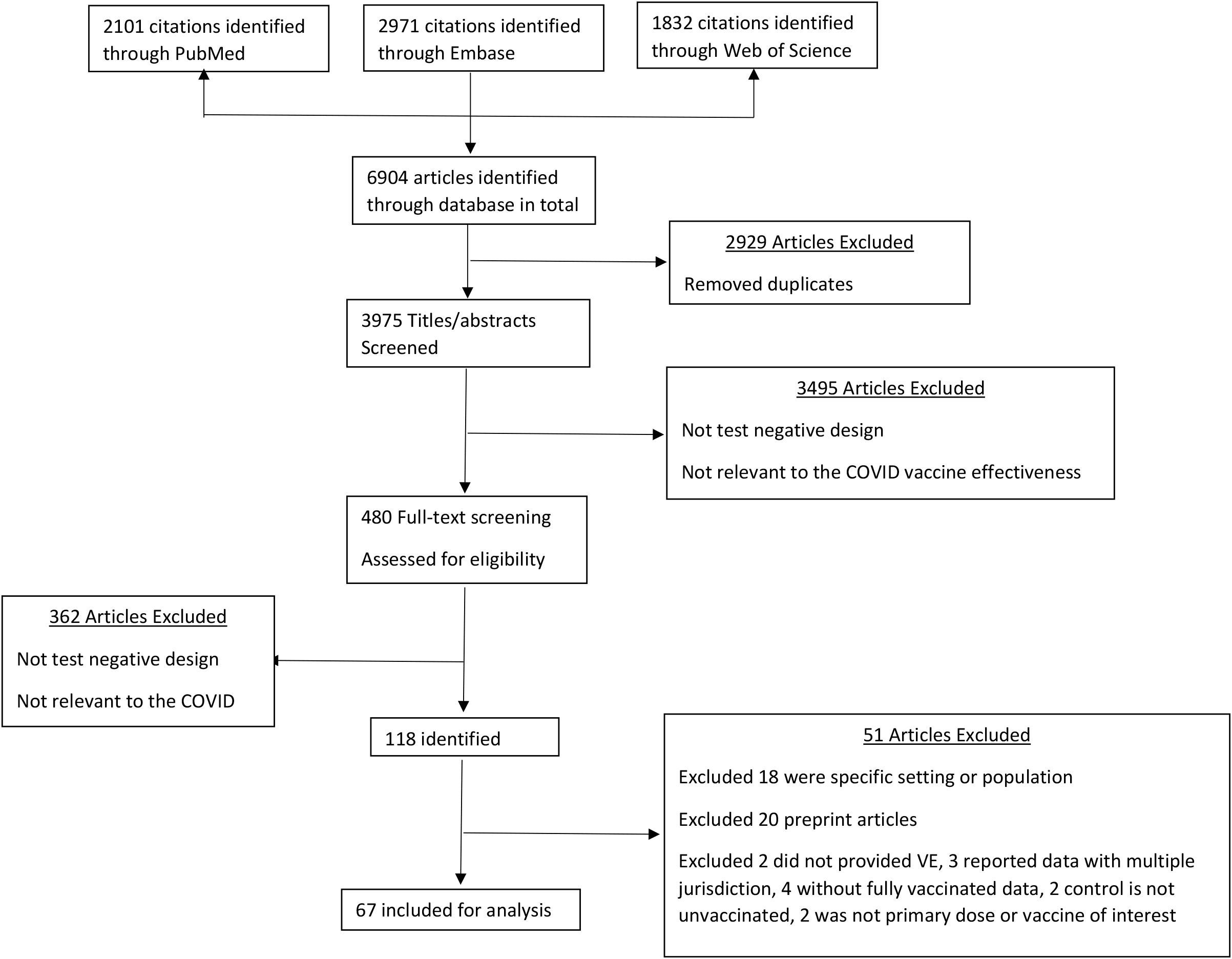
Selection of studies for the systematic review

### Vaccine effectiveness against infection and severe disease

The 173 VE point estimates against infection ranged from 14% to 98%, with I^2^=100%, indicating high heterogeneity (Figure 2-3). Among them, 95 (55%) were higher than 80%. The 93 point estimates against severe disease were also highly heterogeneous (I^2^=100%), ranging from 20% to 100% (Figure 2-3). Among them, 71 (76%) were higher than 80%. For both outcomes, we observed declining VE over time from early 2021 to mid 2022 (Figure S5).

**Figure 2.**
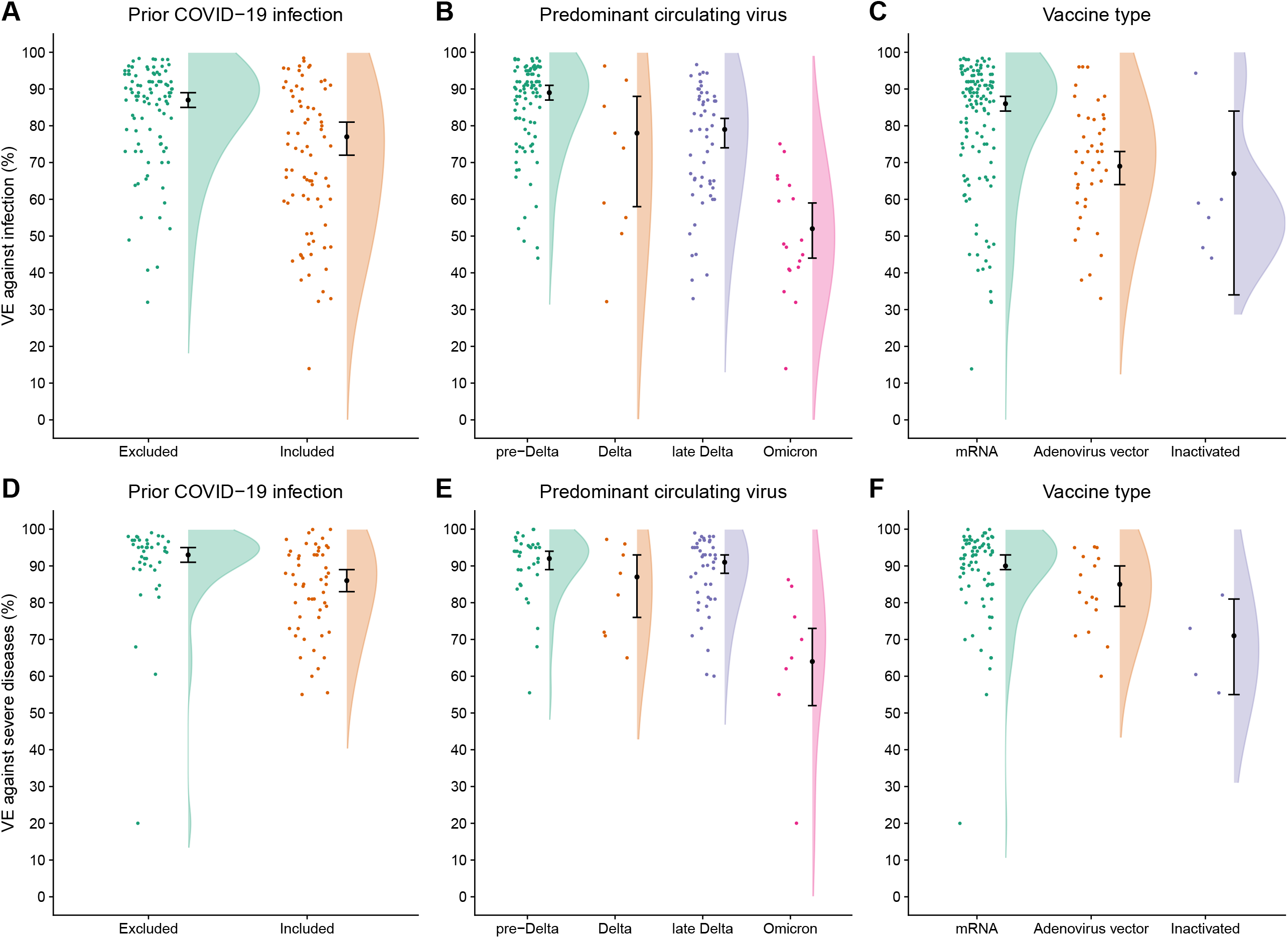
VE point estimates from identified studies based on prior infection (Panel A), predominant circulating virus (Panel B) and vaccine type (Panel C). Each point represents the VE point estimate. Estimates are jittered to enable visualisation. Black points represent the pooled VE estimate from meta-analysis with black lines representing the 95% confidence interval around the pooled estimate. Shaded area is the violin plot, showing the smoothed density of the VE point estimates.

**Figure 3.**
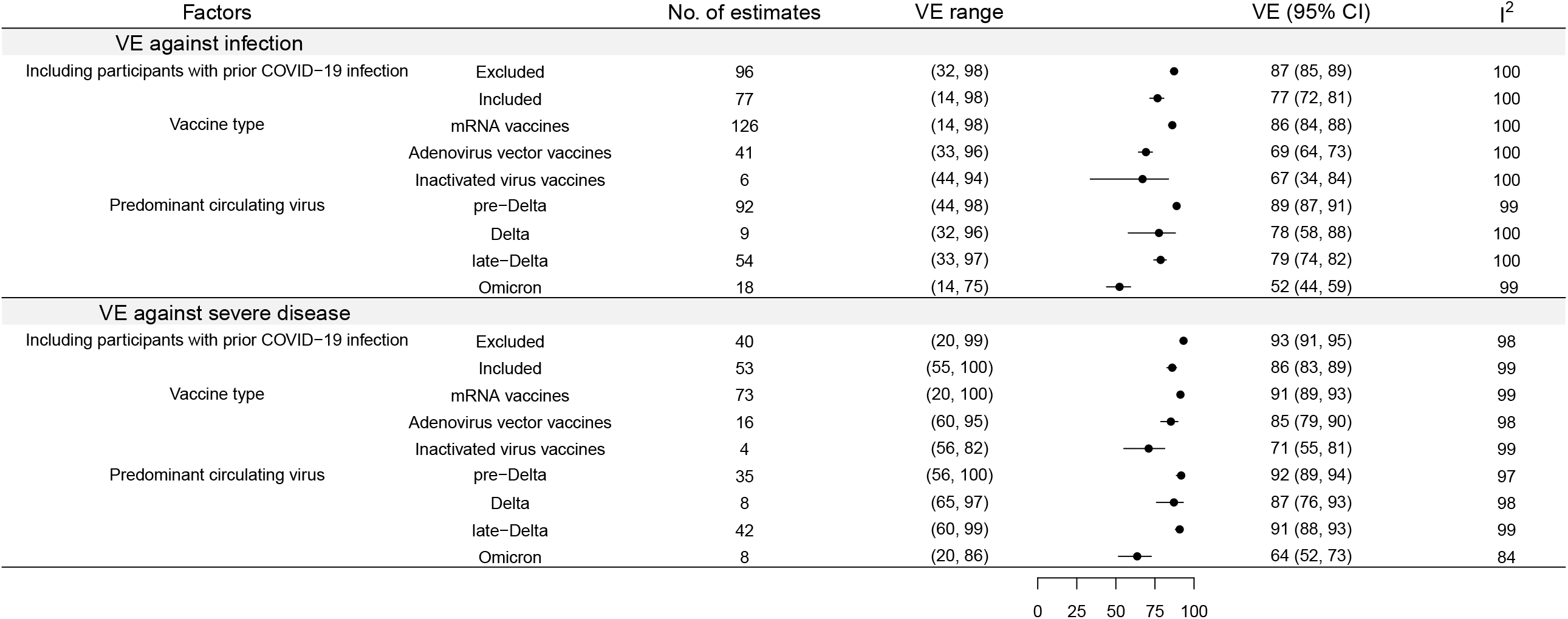
Pooled VE estimates against infection and severe disease by circulating virus, vaccine types, and the inclusion or exclusion of participants with prior COVID-19 infection from random-effect meta-analysis.

### Impact of type of vaccine and circulating viruses

Our meta-analysis (Figure 3) indicated that pooled VE against infection for a primary course of mRNA vaccines was 86% (95% CI: 84%, 88%), compared to 69% (95% CI: 64%, 73%) for adenovirus vector vaccines and 67% (95% CI: 34%, 84%) for inactivated virus vaccines. When we examined differences in pooled VE by the circulating virus, we found that VE against infection during the Omicron period was far lower (VE: 52%; 95% CI: 45%, 59%) than during the pre-Delta (VE: 89%; 95% CI: 87%, 91%), Delta (VE: 78%; 95% CI: 58%, 88%), and the late-Delta periods (VE: 79%; 95% CI: 74%, 92%). Similarly, VE against severe disease during the Omicron period was 64% (95% CI: 56%, 71%), which was lower than for pre-Delta (VE: 92%; 95% CI: 89%, 94%), Delta (VE: 87%; 95% CI: 76%, 93%), and late-Delta periods (VE: 91%; 95% CI: 88%, 93%). The results were similar when further disaggregated by including or excluding prior infection (Table S8), or using fixed-effects analysis (Figure S6).

### Role of prior infection on VE estimates

In general, we found that VE estimates derived from study participants with lower pre-existing immunity were higher. The pooled VE against infection for studies that excluded participants with prior COVID-19 infection was higher (VE: 87%; 95% CI: 85%, 89%) than from studies that included these participants (VE: 77%: 95% CI: 72%, 81%). Similarly, pooled VE against severe disease from studies that excluded participants with prior COVID-19 infection (VE: 93%; 95% CI: 91%, 95%) was higher than from studies that included these participants (VE: 87%: 95% CI: 84%, 90%). There was high heterogeneity among the estimates (I^2^ > 99%). The pooled estimates from fixed-effect analysis were similar (Figure S6).

In meta-regression adjusting for vaccine type, circulating virus, and enrolment criteria (Table S6; Figure 4A-B), the OR against infection from studies that included participants with prior COVID-19 infection (higher pre-existing immunity) was 1.56-fold higher (95% CI: 1.29, 1.89) than the OR from studies that excluded these participants (i.e. with generally lower pre-existing immunity). Therefore, the VE against infection in a study that originally excluded participants with prior COVID-19 infection was 80%, it would be expected to yield an estimate of 69% (95% CI: 62%, 74%) had they included those participants. Similarly, the OR against severe disease from studies that included participants with prior COVID-19 infection was 1.73-fold higher (95% CI: 1.23, 2.45) than from studies that excluded these participants. Assuming a baseline VE against severe disease of 90%, the corresponding VE expected when participants with prior infection were included would be 83% (95% CI: 76%, 87%). The results were similar with adjustment for location and duration of study (Table S7).

**Figure 4.**
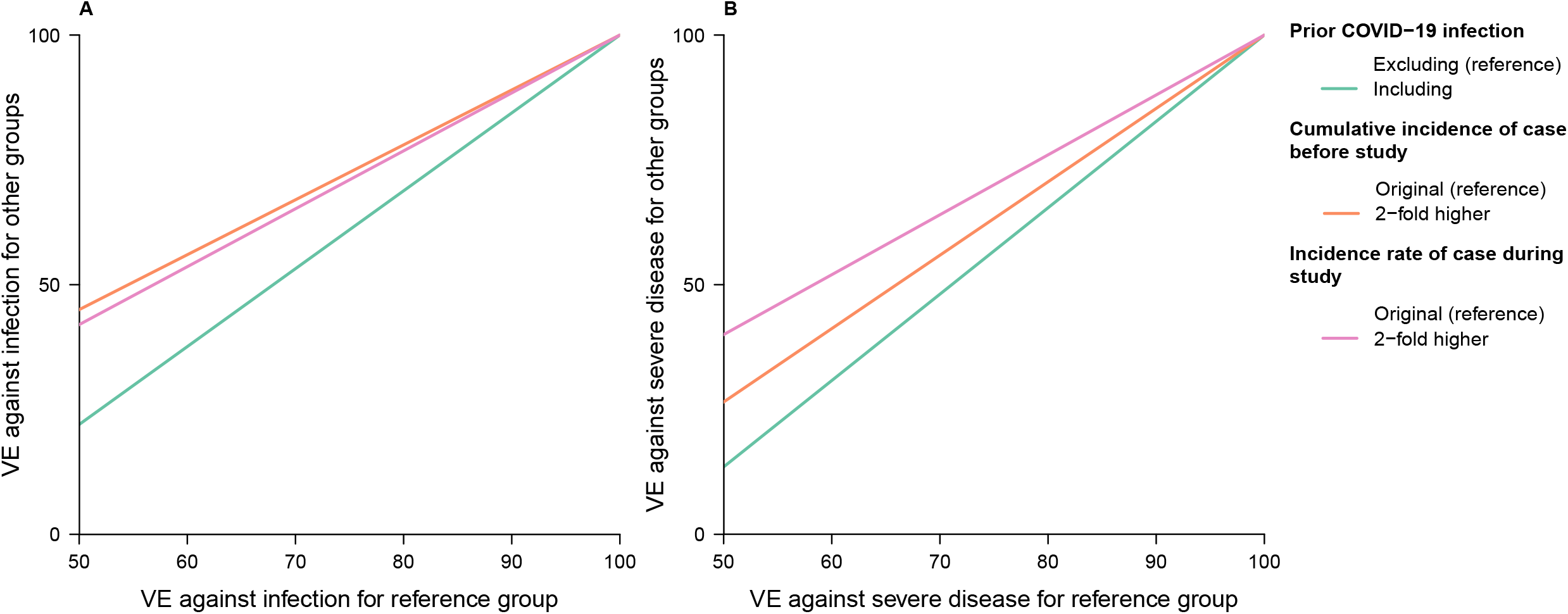
Predicted VE for a group of individuals based on the estimated ratio of odds ratios (ROR) estimated from meta-regression, and the VE for the individuals in the reference group. Panel A and B indicate predicted VE against infection and severe disease, respectively. Prior COVID-19 infection, cumulative incidence of cases before the study, and the incidence rate of cases during study are considered. Models adjusted for age group, type of vaccine, predominant circulating virus, and enrolment criteria.

### Impact of cumulative incidence

There was a modest, negative correlation between the cumulative incidence of cases in the study locations prior to the start of the study, as a proxy of pre-existing population immunity (Figure S8), and VE against infection (Pearson correlation (*r*) = −0.42; 95% CI: −0.54, −0.30; Shearman correlation (*ρ*) = −0.32; 95% CI: −0.45, −0.18) and severe disease (*r* = −0.41, 95% CI: −0.56, −0.22; *ρ* = −0.49; 95% CI: −0.64, −0.30). In meta-regression, adjusting for vaccine type, circulating virus, and enrolment criteria (Table S6; Figure 4A-B), the ROR against infection associated with a doubling of the cumulative incidence of cases before the start of studies was 1.10 (95% CI: 1.02, 1.20). Therefore, if the baseline VE against infection from a study was 80%, then the corresponding VE for a setting with twice the cumulative incidence of cases before the start of a study would represent a two-percentage point reduction in VE (VE=78%; 95% CI: 76%, 79.6% for a doubling). The ROR against severe disease for each doubling of the cumulative incidence of cases before the start of a study (higher pre-existing immunity) was 1.47 (95% CI: 1.26, 1.71). Therefore, assuming a baseline VE against severe diseases of 90%, the corresponding VE for a setting with twice the cumulative incidence of cases before the start of a study would represent a 5 percentage point drop in VE (VE=85%, 95% CI: 83%, 87% for the initial doubling).

### Impact of incidence rate during the study period

There was a modest, negative correlation between the incidence rates of cases in the study locations prior to the start of the study, as a proxy of pre-existing population immunity (Figure S8), and VE against infection (*r* = −0.38; 95% CI: - 0.50, −0.24; *ρ* = −0.48; 95% CI: −0.59, −0.34) and severe disease (*r* = −0.50, 95% CI: −0.64, −0.33; *ρ* = −0.42; 95% CI: −0.58, −0.23). After adjusting for vaccine type, circulating virus and enrolment criteria (Table S6), we estimated that the ROR against infection for each doubling of the incidence of cases during the study period was 1.16 (95% CI: 1.07, 1.25). If the baseline VE against infection from a study was 80%, then the corresponding VE from a study with twice the incidence of cases during the study period would be 77% (95% CI: 75%, 79%). We also estimated that the ROR against severe disease associated with a doubling in the incidence of cases and death during the study was 1.20 (95% CI: 1.03, 1.40). Therefore, assuming a baseline VE of 90% against severe disease, the corresponding VE for a study with twice as many cases during the study period would be 88% (95% CI: 86%, 89.7%).

## DISCUSSION

In this study, we summarized VE estimates from TND studies to understand the impact of prior infections on VE estimates. We found that higher pre-existing immunity in the source population, indicated by including participants with prior COVID-19 infection, higher pre-study cumulative incidence of cases, and higher incidence rate of cases during study period, was associated with lower VE.

Prior infection could be a confounder, effect modifier or both. As a confounder it could affect peoples’ decisions to vaccinate and modify their risk behaviours as well as providing protection against reinfection (12, 13, 21). Hence, the VE obtained from individuals with or without prior infection would be similar, if the influence of the confounding could be controlled in analysis. On the other hand, if prior infection were only an effect modifier (i.e. only associated with the risk of (re)infection and not the propensity to be vaccinated) vaccination in settings with higher pre-existing immunity would appear to have a relatively modest effect on further increasing protection at the population level because VE would be lower among previously infected participants (12, 13). In reality prior infection is probably both a confounder and effect modifier and therefore studies should consider both appropriate confounding control, such as through covariate adjustment or stratification, as well as inclusion of interaction terms to explore the potential effect modification.

When VE was estimated based on studies excluding participants with prior infection, these VE estimates should be interpreted as the VE for a hypothetical population with no pre-existing immunity. As of late 2022, these estimates would have limited practical value in most locations which have experienced substantial epidemics. Epidemic forecasting models used to inform public health control policies should separate individuals into different compartments based on infection history to improve the precision of their forecasts. Therefore, groups estimating VE estimates to inform policy should stratify by infection history so that their work will be more broadly useful for policy (100, 101).

Our observation that higher incidence rates during a study period were associated with lower VE estimates suggests that SARS-CoV-2 vaccines provide leaky protection (102), since the VE depended on the number of exposure (proxied by incidence rates during study period). It has previously been shown that the ORs derived from TND studies were biased (103), so that VE estimates for leaky vaccines would decrease with time since vaccination, even if the true VE remained unchanged (102). For COVID vaccines, the antigenic drift observed for SARS-CoV-2 viruses makes it difficult to disentangle reduced VE associated with a leaky vaccine from reduced VE associated with vaccine antigenic match, and a decreased proportion of susceptible in the community.

Although 55% of VE estimates against infection and 76% of estimates against severe disease were higher than 80%, heterogeneity was very high, as indicated by the high I^2^ values observed. Consistent with previous reviews, high heterogeneity could be attributed to differences in effectiveness among vaccine types, or the predominant circulating virus in each study (8, 105). However, we continued to observe high heterogeneity when estimating pooled VE against specific vaccine types and the predominant virus. Our meta-regression identified some sources of the heterogeneity, such as pre-existing immunity. However, heterogeneity remained high and further investigation is needed to identify other causes to ensure valid VE estimates are available for ongoing optimization of vaccination strategies (106).

Our study had some limitations. First, our review focused on VE of primary vaccination series. Further analysis would be required to determine whether similar issues apply to estimation of VE for booster doses, which are complicated by dosing schedules that mix vaccine types, the number of doses received, greater antigenic differences between the vaccines received and the dominant circulating virus, changes in vaccine formulation including bivalent formulations, and the accumulation of immunity through both vaccination and infection over time. Second, most studies were conducted in adults, so that our results may not be generalizable to children. Finally, TND studies included in our review were observational in nature. Some confounders were adjusted in these studies, including age, sex, being health care workers, or pre-existing conditions. However, we did not include a bias assessment to evaluate whether studies adequately addressed confounding nor have we considered other potential sources of bias such as measurement errors.

In conclusion, we observed reduced VE associated with higher pre-existing immunity in the population. Exclusion of participants with prior infection could artificially inflate VE estimates and affect their generalisability to the wider population. If the goal of a study is to inform policy that applies to the whole population, participants with prior infection should be included and their status included as a covariate for confounder control. However, if decision-makers desire different vaccination policies dependent on infection history then studies need to stratify accordingly, or including interaction term, instead of excluding participants with prior infection. Studies unable to adjust for prior infection could consider using external adjustment (107) to assess the potential effect of this confounder on their estimates. Optimal design of VE studies remains a research priority. In particular, further work is needed to understand how prior infection influences VE for booster doses and as vaccine formulations change.

## Supporting information

Supplemental appendix, Supplemental research in context

## Data Availability

All data produced in the present work are contained in the manuscript

## ACKNOWLEDGEMENTS

The authors thank Hang Qi for technical assistance. This project was supported by the National Institute of General Medical Sciences (grant no. R01 GM139926), and the Theme-based Research Scheme (Project No. T11-712/19-N) of the Research Grants Council of the Hong Kong SAR Government. BJC is supported by an RGC Senior Research Fellowship (grant number: HKU SRFS2021-7S03) and the AIR@innoHK program of the Innovation and Technology Commission of the Hong Kong SAR Government. The WHO Collaborating Centre for Reference and Research on Influenza is supported by the Australian Government Department of Health and Aged Care.

