## Supplemental appendix, Supplemental research in context for "Prior infections and effectiveness of SARS-CoV-2 vaccine in test-negative study: A systematic review and meta-analysis": Research_in_context.docx

**Evidence before this study**

Accurate estimate of vaccine effectiveness (VE) of COVID-19 is important to guide control policy. Multiple studies have been conducted to evaluate and monitor VE, particularly the test-negative design (TND) studies. Pre-existing population immunity caused by infections could have impact on VE of COVID-19 vaccine, which may also contribute to the variation of VE estimates from different studies. A search of PubMed on 2 November 2022, using search term describing VE against SARS-CoV-2 infection and severe disease with no language restrictions identified 30 systematic reviews on the VE of COVID-19 vaccine, including VE against infection, symptomatic cases, severe diseases, or fatality. These studies focus on the VE in different groups and settings, including variants, age groups, type of vaccines, and people with pre-existing conditions, or specific groups of individuals (e.g. health care worker). However, none of the reviews examined the impact of pre-existing immunity on the VE estimates from vaccine trials, or TND studies.

**Added value of this study**

To fill this research gap, we did a systematic review and meta-analysis of TND studies estimating VE against infection and severe disease, based on their inclusion criteria (including or excluding individuals with prior infection). We systematically searched PubMed, Embase and Web of Science on 11 July 2022. We examined the impact of pre-existing immunity on VE estimates, using 1) including or excluding individuals with prior infection, 2) the cumulative incidence of cases prior to the study in the study region, and 3) the incidence rate of cases during the study in the study region.

To our knowledge, this is the first systematic review and meta-analysis examining the impact of pre-existing immunity on VE estimates. We identified 67 TND studies. Then, we conducted meta-analysis and found that the pooled VE among studies that included people with prior infection was lower, compared with studies that excluded people with prior COVID-19 infection. We also estimated a negative correlation between cumulative incidence of cases before the start of study and VE estimates. The same negative correlation was also observed for incidence rate of case during study period and VE estimates. In the meta-regressions that adjusted for age group, type of vaccine, predominant circulating virus and enrolment criteria, we still observed the same association. Our review suggested that higher level of pre-existing immunity in a population was associated with low VE estimates.

**Implications of all the available evidence**

Excluding participants with prior infection may inflate VE estimates and affect the generalizability to the wider population. Prior infection could be a confounder, effect modifier or both. Participants with prior infection should be included, and the prior infection status could be considered as a covariate for confounder control, if the VE estimates are used to inform control policy for the whole population. When vaccination policy is decided to be dependent of infection history, then studies may need to stratify, or including interaction term, to obtain the corresponding VE estimates for individuals with or without prior infections. Optimal design of VE studies remains a research priority.
