## Supplemental appendix, Supplemental research in context for "Prior infections and effectiveness of SARS-CoV-2 vaccine in test-negative study: A systematic review and meta-analysis": TND_appendix_v10.docx

**Supplementary Note 1. Directed cycled graph to demonstrate the role of prior infection in estimation of vaccine effectiveness.**

Prior infection

Vaccination	Immunity	COVID-19

Prior infection could act as a confounder because previous infection may influence people’s decision to be vaccinated and may provide latent protection against infection.

Prior infection

Vaccination	Immunity	COVID-19

Prior infection could act as an effect modifier, one example is:

Previously infected people may have acquired some immunity other than antibody, such as cell-mediated immunity, which may have interaction of the protection from vaccination.

It should be noted that these two DAG are just used to show that prior infection could act as a confounder and an effect modifier, they are not mutually exclusive, such as the following DAG:

Prior infection

Vaccination	Immunity	COVID-19

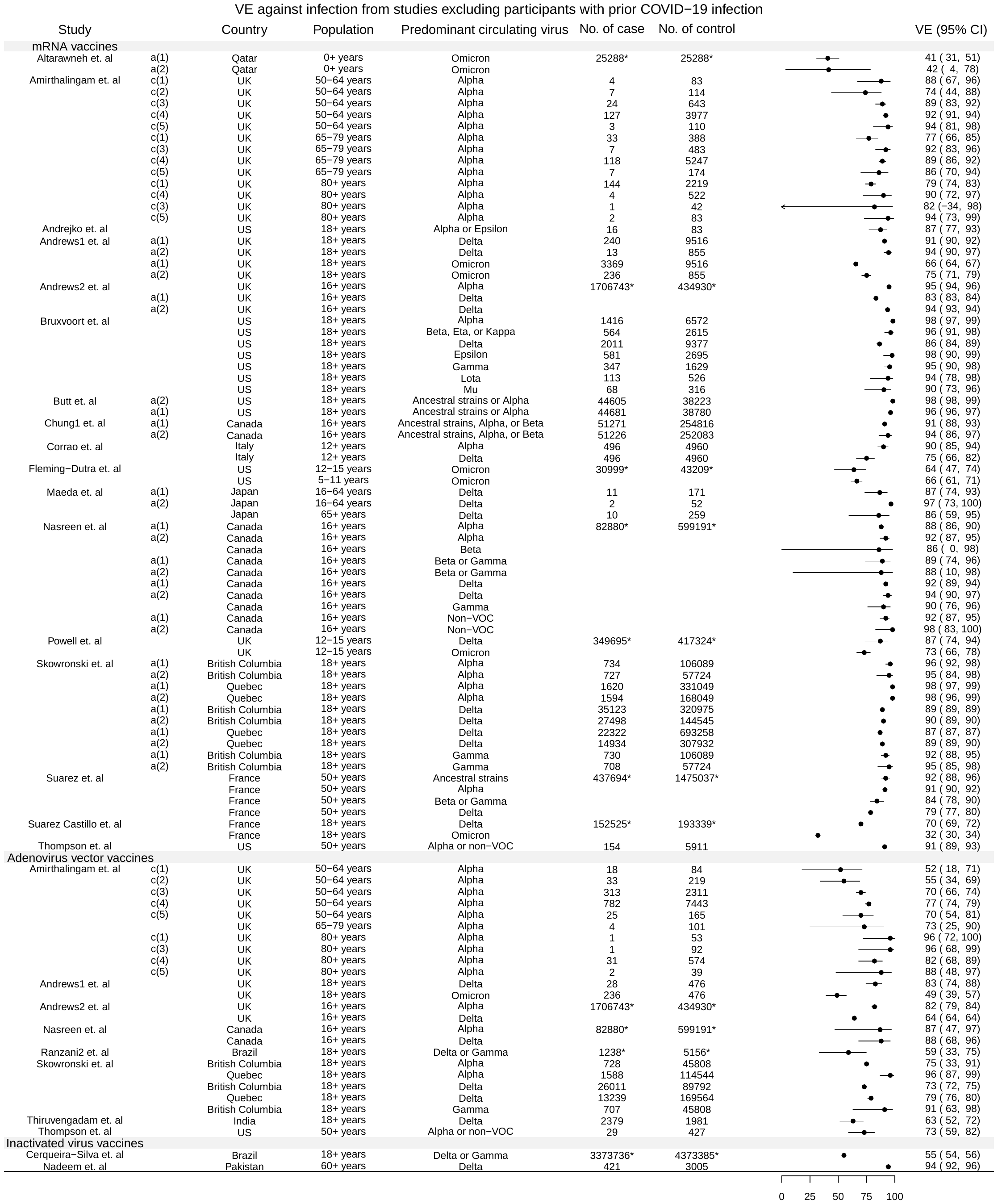

**Figure S1.** Estimates of VE against infection from identified studies that excluded participants with COVID-19 infection history. Multiple estimates could be due to: a. vaccine, (1) for PZ, (2) for Moderna, (3) for ChAdOx1, (4) for rad26-rad5(Sputnik V); b. control: (1) syndrome-negative, (2) test-negative control; c. different duration between first and second dose, (1) for 19-29 days, (2) for 30-44 days, (3) for 45-64 days, (4) for 65-84 days, (5) for 85+ days

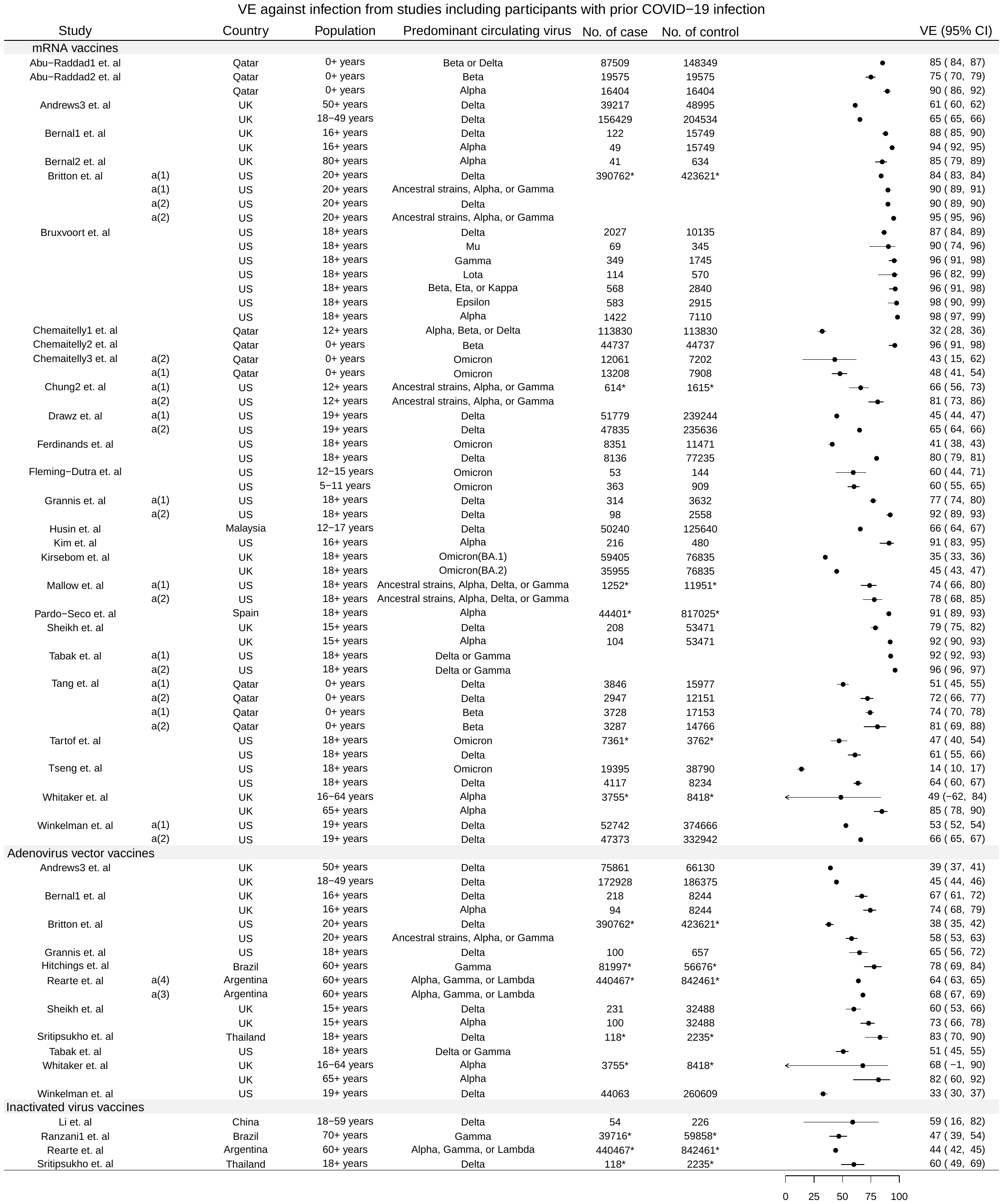

**Figure S2.** Estimates of VE against infection from identified studies that included participants with COVID-19 infection history. Multiple estimates could be due to: a. vaccine, (1) for PZ, (2) for Moderna, (3) for ChAdOx1, (4) for rad26-rad5(Sputnik V); b. control: (1) syndrome-negative, (2) test-negative control; c. different duration between first and second dose, (1) for 19-29 days, (2) for 30-44 days, (3) for 45-64 days, (4) for 65-84 days, (5) for 85+ days

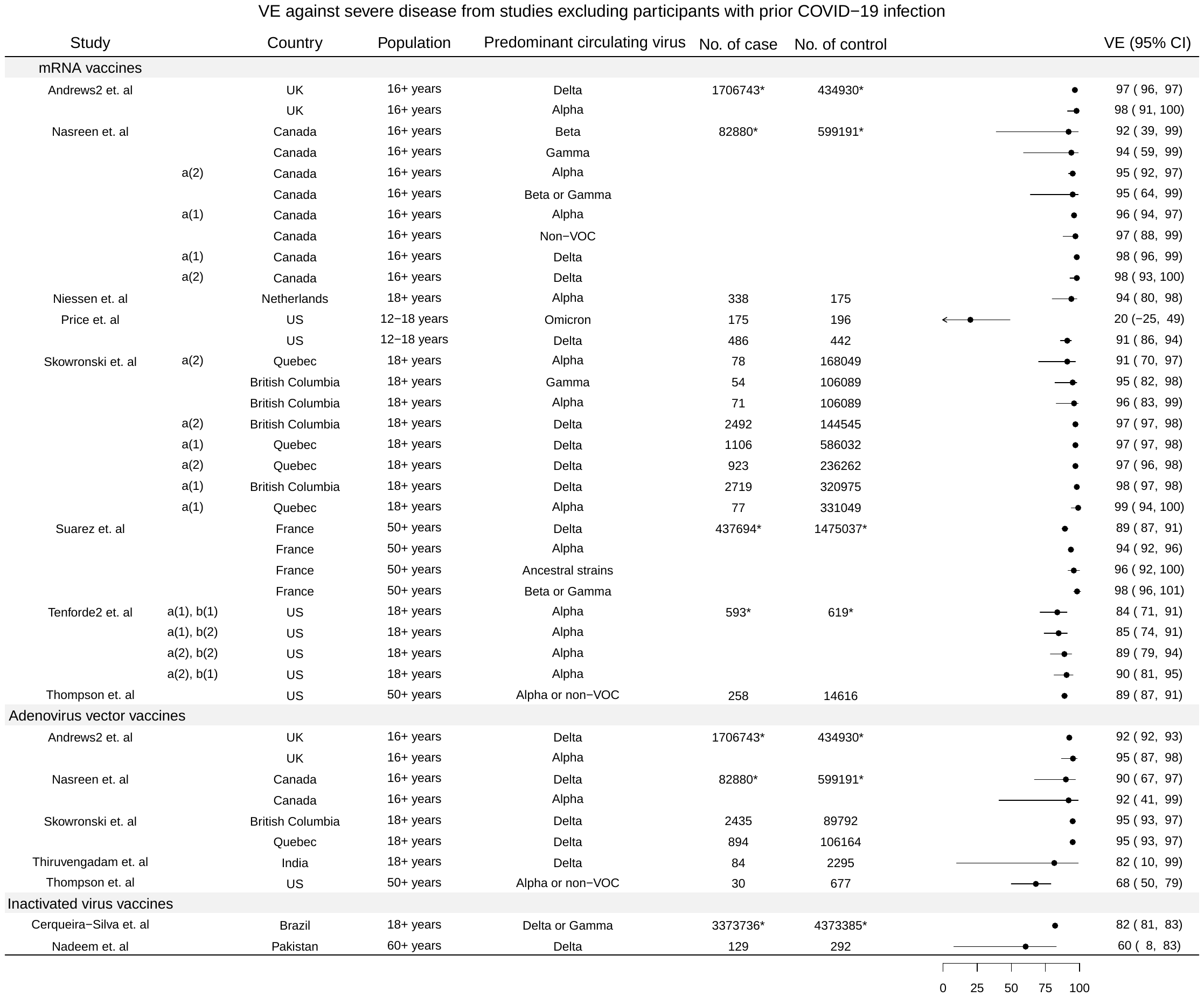
**Figure S3.** Estimates of VE against severe disease from identified studies that excluded participants with COVID-19 infection history. Multiple estimates could be due to: a. vaccine, (1) for PZ, (2) for Moderna, (3) for ChAdOx1, (4) for rad26-rad5(Sputnik V); b. control: (1) syndrome-negative, (2) test-negative control; c. different duration between first and second dose, (1) for 19-29 days, (2) for 30-44 days, (3) for 45-64 days, (4) for 65-84 days, (5) for 85+ days

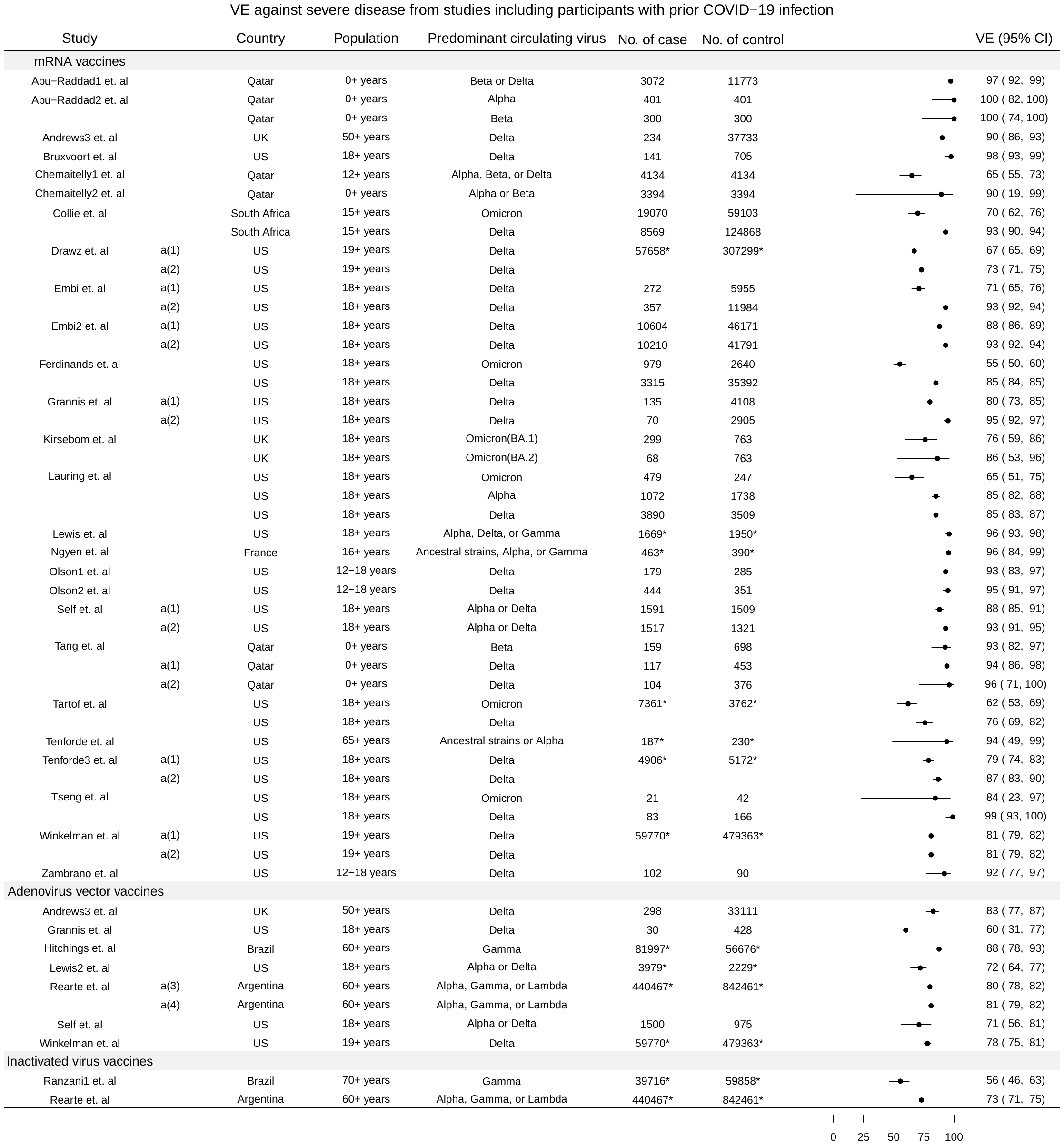

**Figure S4.** Estimates of VE against severe disease from identified studies that included participants with COVID-19 infection history.

Multiple estimates could be due to: a. vaccine, (1) for PZ, (2) for Moderna, (3) for ChAdOx1, (4) for rad26-rad5(Sputnik V); b. control: (1) syndrome-negative, (2) test-negative control; c. different duration between first and second dose, (1) for 19-29 days, (2) for 30-44 days, (3) for 45-64 days, (4) for 65-84 days, (5) for 85+ days

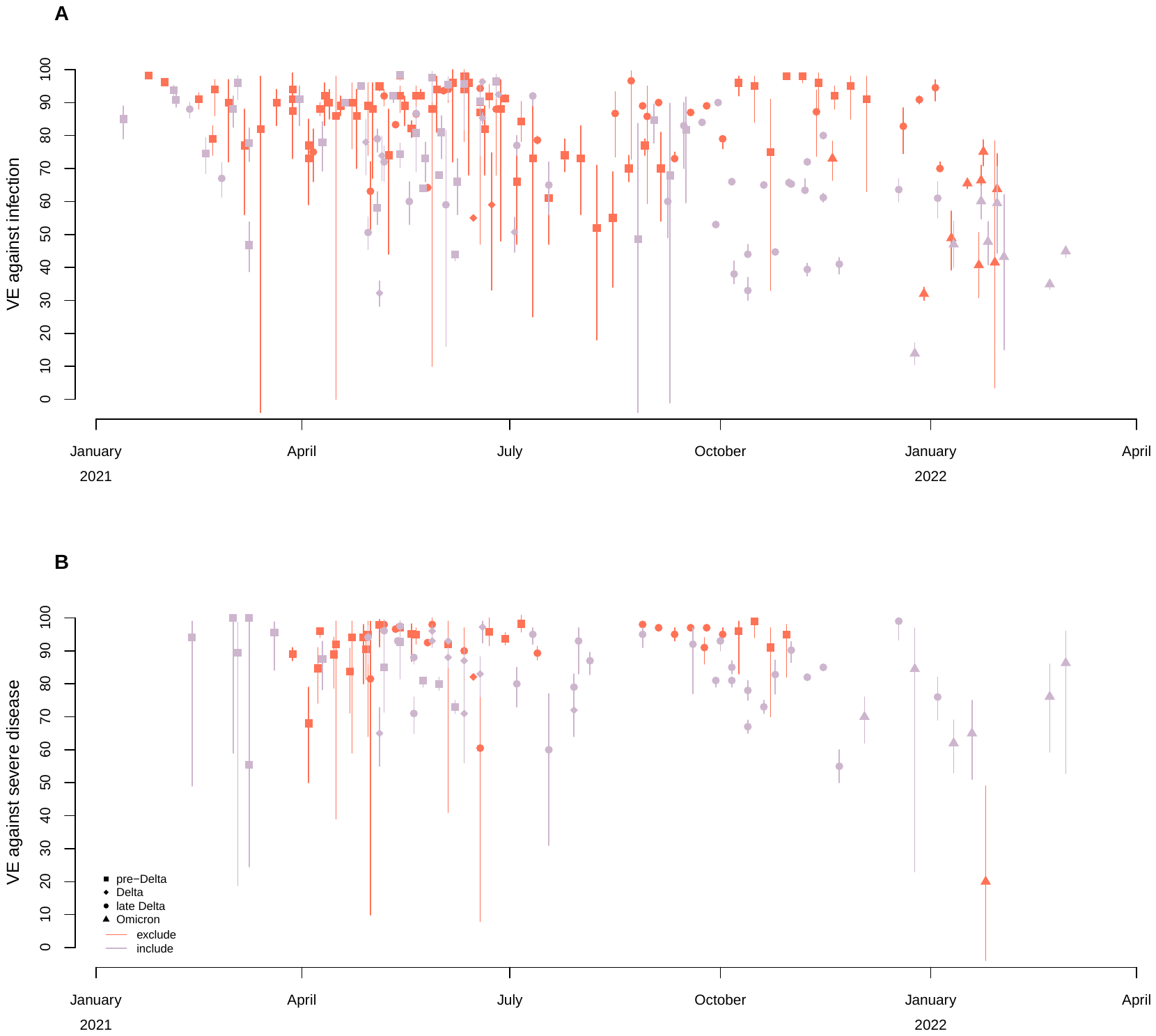

**Figure S5.** VE estimates extracted from studies, by calendar time.

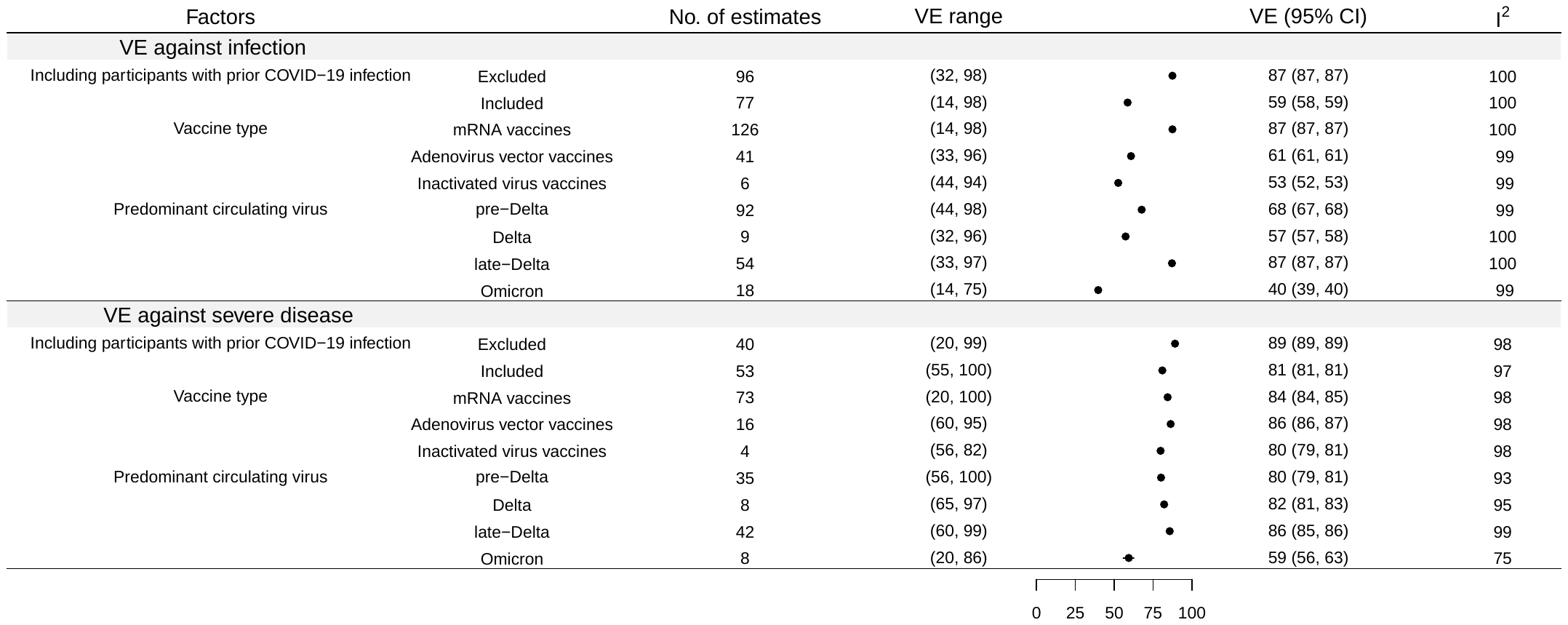

**Figure S6.** Pooled VE estimates against infection and severe disease by circulating virus, vaccine types, and the inclusion or exclusion of participants with prior COVID-19 infection, from fixed-effect meta-analysis.

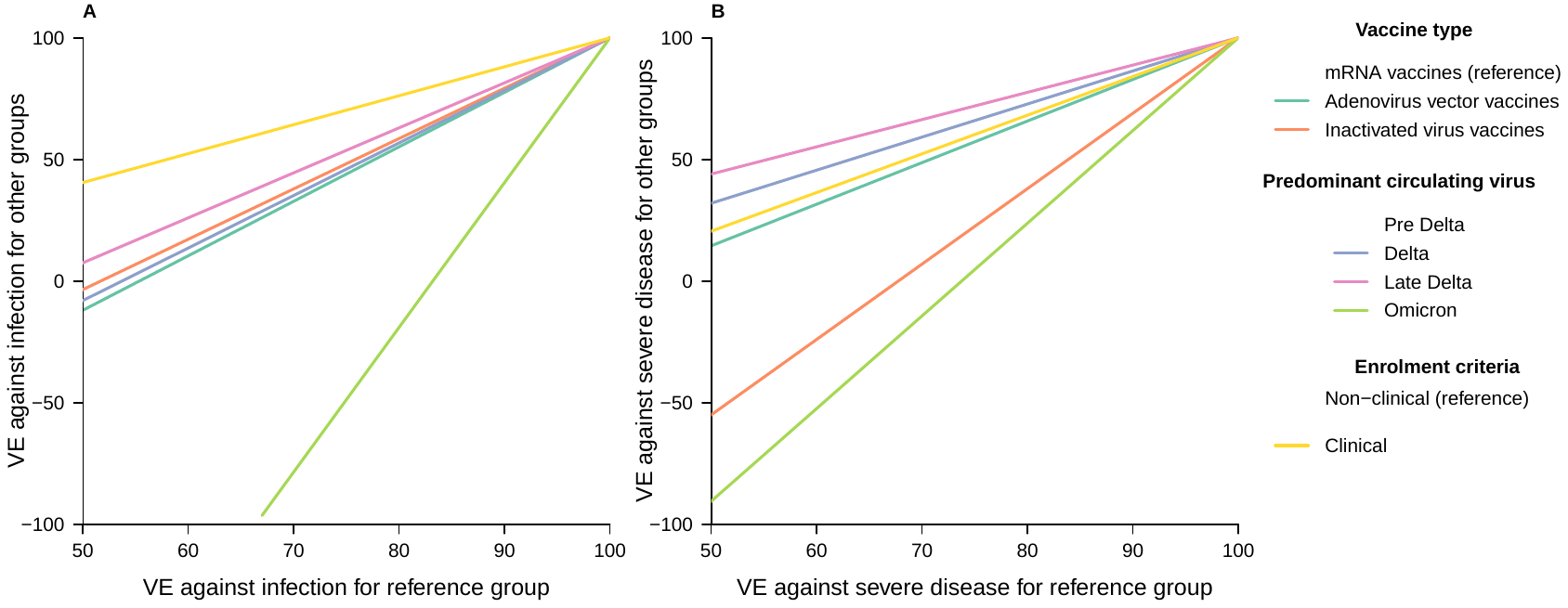

**Figure 4.** Predicted VE for a group of individuals based on the estimated ratio of odds ratio (ROR) estimated from meta-regression, and the VE for the individual in reference group. Panel A and B indicate predicted VE against infection and severe disease respectively. Vaccine type, predominant circulating virus and enrolment criteria are considered.

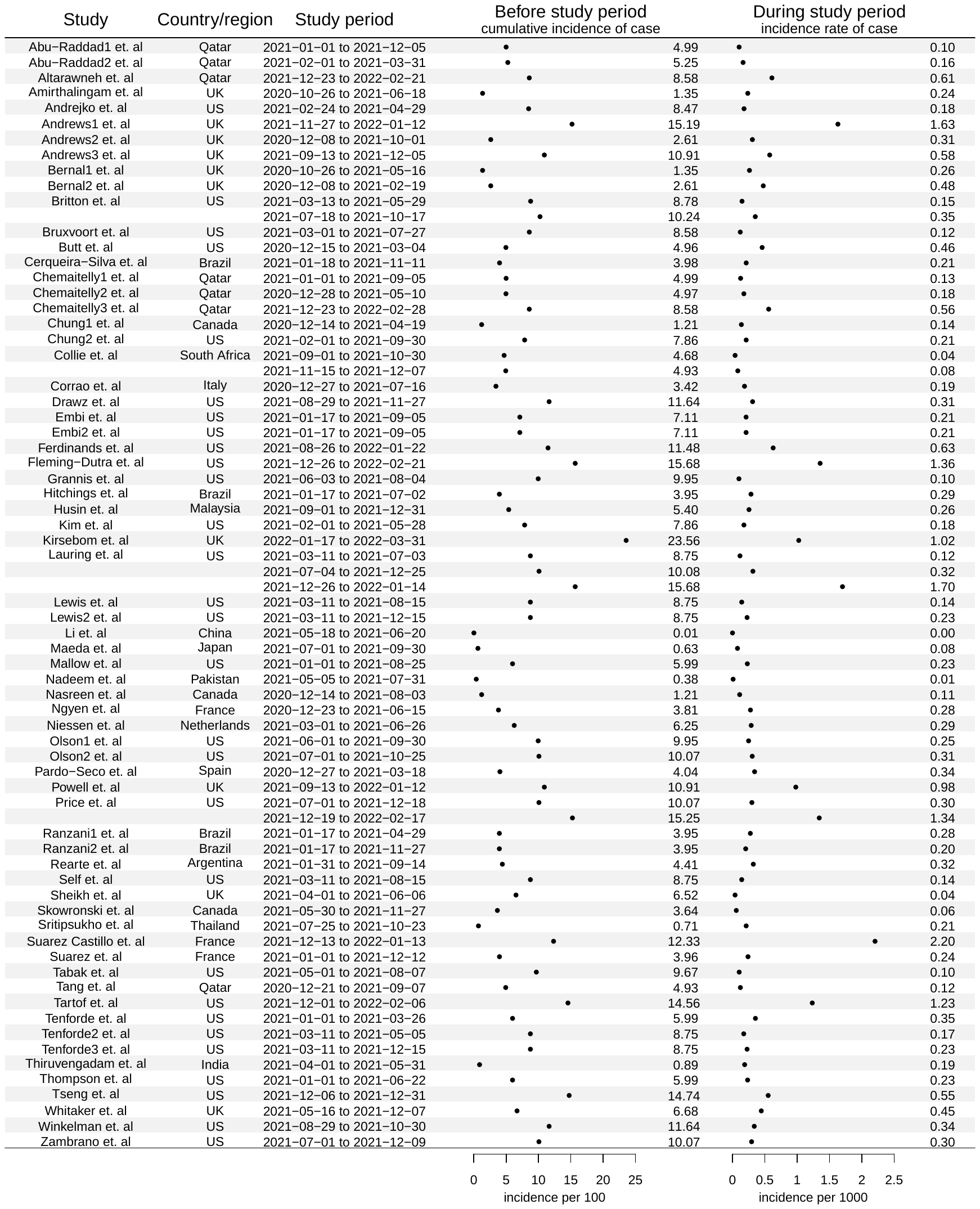

**Figure S8:** the cumulative incidence of case before the study period and incidence rate of case during study for each identified study.

**Table S1.** Variables in extraction form

| variable | description |
| --- | --- |
| study id | Identity number of the study |
| author | first author of the study |
| year | publish year |
| paper | published paper |
| pmid | pmid o the study |
| study type | general type of the study |
| study | specific type of study |
| study_period_start | the start time point of the study period |
| study_period_end | the end time point of the study period |
| vaccine | vaccine type used |
| historyofCOVID | how to dealing with those with covid history, included(0): included those with covid history, excluded(1): excluded those with covid history |
| dose_number | total dose number of the vaccine |
| dose | dose number |
| timing_of_dose_days | the time after vaccination |
| outcome | endpoint of the study |
| country | study location |
| population | study population |
| VE | vaccine effectiveness |
| LCI | the lower bound of 95% confidence interval for vaccine effectiveness |
| UCI | the upper bound of 95% confidence interval for vaccine effectiveness |
| no of case(total) | total number of cases |
| no of case(vac) | total number of vaccinated cases |
| no of case(unvac) | total number of unvaccinated cases |
| no of control(total) | total number of control |
| no of control(vac) | total number of vaccinated control |
| no of control(unvac) | total number of unvaccinated control |
| variant | predominant circulating virus during the study period |
| overall | for overall/individual vaccine estimate, NA:that study reported either overall vaccine or individual vaccine estimate; 1:reported both overall vaccine and individual vaccine estimate, and the corresponding row is for overall vaccine; 0:reported both overall vaccine and individual vaccine estimate, and the corresponding row is for individual vaccine |
| adjust for | adjustment of the estimate |
| adjust | the estimate provided is adjusted or not, 1:with adjustment, 0: without adjustment |
| by_clinial | enrolment critera, 1: clinical, 0: non-clinical |

**Table S2**. Summary of included studies in the systematic review and meta-analysis

| **Author (year)** | **Participant** | **Study period** | **Location** | **COVID-19 infection history** | **Vaccine type** | **Circulating Virus type** | **Endpoint type** | **Recruitment criteria** | **Adjustments in analysis** |
| --- | --- | --- | --- | --- | --- | --- | --- | --- | --- |
| Abu-Raddad1 (2022) | general | 2021-01-01 to 2021-12-05 | Qatar | Included | mRNA | Beta & delta | Infection & severe | Non-clinical | Prior infection, healthcare worker status |
| Abu-Raddad2 (2021) | general | 2021-02-01 to 2021-03-31 | Qatar | Included | mRNA | Alpha & Beta & other variants | Infection & severe | Non-clinical | Sex, age, nationality, PCR test date |
| Altarawneh  (2022) | general | 2021-12-23to2022-02-21 | Qatar | Excluded | mRNA | Omicron | Symptomatic | Non-clinical | Sex, 10-year age group, nationality, and calendar week of PCR test |
| Amirthalingam (2021) | $\geq$50 years of age | 2020-10-26 to 2021-06-18 | UK | Excluded | mRNA & Adenovirus vector vaccines | Alpha | Infection | Clinical | Week of onset, 5-year age bands, gender, NHS region, index of multiple deprivation (quintiles), ethnicity, health/social care worker, care home resident |
| Andrejko (2021) | $\geq$18 years of age | 2021-02-24 to 2021-04-29 | US | Excluded | mRNA | Epsilon & alpha | Infection & severe | Non-clinical | Age, region, sex, income, and race predicted the likelihood an individual was vaccine hesitant |
| Andrews1 (2022) | $\geq$18 years of age | 2021-11-27 to 2022-01-12 | UK | Excluded | mRNA & Adenovirus vector vaccines | Delta & omicron | Symptomatic | Clinical | Age (18 to 89 years in 5-year bands, then everyone ≥90 years), sex, index of multiple deprivation (quintile), race or ethnic group, history of foreign travel, geographic region, period (day of test), health and social care worker status, clinical riskgroup status, status of being in a clinically extremely vulnerable group, and previously testing posit |
| Andrews2 (2022) | $\geq$16 years of age | 2020-12-08 to 2021-10-01 | UK | Excluded | mRNA & Adenovirus vector vaccines | Alpha & delta | Infection & severe | Clinical | Age, sex, index of multiple deprivation (a measure of socioeconomic status), race or ethnic group, care home residence status (for analyses including persons ≥65 years of age), geographic region, period (calendar week), health and social care worker status (for analyses involving persons <65 years of age), and status of being in a clinical risk group (available only for persons <65 years of age) or a clinically extremely vulnerable group (any age) |
| Andrews3  (2022) | $\geq$18 years of age | 2021-09-13to2021-12-05 | UK | Included | mRNA & Adenovirus vector vaccines | Delta | Symptomatic & severe | Clinical | Age (5-year bands), sex, index of multiple deprivation (quintile), ethnic group, care-home residence status, geographic region (NHS region), period (calendar week of onset), health and social care worker status, clinical risk group status, clinically extremely vulnerable, severely immunosuppressed and previously testing positive |
| Bernal1 (2021) | $\geq$16 years of age | 2020-10-26 to 2021-05-16 | UK | Included | mRNA & Adenovirus vector vaccines | Alpha & delta | Symptomatic | Clinical | Period (calendar week), travel history, race or ethnic group, sex, age, index of multiple deprivation, clinically extremely vulnerable group, region, history of positive test, health or social care worker, and care home residence |
| Bernal2 (2021) | $\geq$70 years of age & $\geq$80 years of age | 2020-12-08 to 2021-02-19 | UK | Included | mRNA & Adenovirus vector vaccines | Alpha | Symptomatic | Clinical | Age, period, sex, region, ethnicity, care home, and index of multiple deprivation fifth |
| Britton (2022) | $\geq$20 years of age | 2021-03-13 to 2021-10-17 | US | Included | mRNA | Delta & mix | Symptomatic | Clinical | Age group, race, ethnicity, sex, testing site state, testing site census tract SVI, and calendar date |
| Bruxvoort (2021) | $\geq$18 years of age | 2021-03-01 to 2021-07-27 | US | Included & excluded | mRNA | Alpha & delta & epsilon & gamma & lota & mu & beta & eta & kappa & other variants | Infection | Non-clinical | BMI, smoking, Charlson comorbidity score, frailty index, lung disease, liver disease, kidney disease, immunocompromised status, pregnancy, heart disease, history of COVID-19 infection, number of outpatient and virtual visits, number of ED visits, number of hospitalizations, preventive care, Medicaid, medical center area, KPSC physician/employee status, month of specimen collection, specimen type |
| Butt (2021) | general | 2020-12-15 to 2021-03-04 | US | Excluded | mRNA | Wild type & alpha | Infection | Non-clinical | Age, sex, race, body mass index, Charlson Comorbidity Index score, and geographic location |
| Cerqueira-Silva (2022) | $\geq$18 years of age | 2021-01-18 to 2021-11-11 | Brazil | Excluded | Inactivated virus vaccines | Gamma & delta | Infection & severe | Clinical | Age, sex, temporal trends, state of residence, previous infection,  pregnancy, postpartum period and comorbidities |
| Chemaitelly1 (2021) | general | 2021-01-01 to 2021-09-05 | Qatar | Included & excluded | mRNA | Alpha & beta & delta | Infection & severe | Non-clinical | Prior infection & healthcare worker status |
| Chemaitelly2 (2021) | general | 2020-12-28 to 2021-05-10 | Qatar | Included | mRNA | Alpha & beta & mix | Infection & severe | Non-clinical | Sex, age, nationality, reason for PCR testing and calendar week |
| Chemaitelly3  (2022) | general | 2021-12-23to2022-02-28 | Qatar | Included | mRNA | Omicron | Symptomatic | Non-clinical | Sex, 10-year-age group, nationality, and calendar week of PCR test |
| Chung1 (2021) | $\geq$16 years of age | 2020-12-14 to 2021-04-19 | Canada | Excluded | mRNA | Wild type & alpha & beta | Symptomatic & severe | Clinical | Age, sex, public health unit region, biweekly period of test, number of SARS-CoV-2 tests in the 3 months prior to 14 December 2020, presence of any comorbidity that increase the risk of severe COVID-19, receipt of influenza vaccination in current or prior influenza season, and neighborhood-level household income, persons per dwelling, proportion of persons employed as non-health essential workers, and self-identified visible minority quintiles |
| Chung2 (2022) | $\geq$12 years of age | 2021-02-01 to 2021-09-30 | US | Included | mRNA | Mix | Symptomatic | Clinical | Study site, age in years (continuous), enrollment period, and self-reported race & ethnicity |
| Collie (2022) | general | 2021-09-01 to 2021-12-07 | South Africa | Included | mRNA | Delta & omicron | Severe | Non-clinical | Age, sex, previous Covid-19 infection, surveillance week, geographic location, and the number of CDC risk factors |
| Corrao (2022) | general | 2020-12-27 to 2021-07-16 | Italy | Excluded | mRNA | Alpha & delta | Infection & severe | Non-clinical | The number of previous contacts with the Regional Health Service, use of corticosteroids, drugs for chronic pain, oral anticoagulant agents and insulin, and the presence of anaemias, chronic respiratory disease, dyslipidaemia, depression, hypertension, coronary and peripheral vascular disease, hypothyroidism, epilepsy and recurrent seizures, psychosis, diabetes without insulin therapy, malignancies, other diseases of the respiratory system, other diseases of the digestive system, other diseases of the genitourinary system, gout, autoimmune disease, other diseases of the circulatory system, symptoms, signs and ill-defned conditions, diseases of the skin and subcutaneous tissues, arrhythmia, infammatory bowel diseases, other mental disorders, heart failure, glaucoma and chronic kidney disease |
| Drawz (2022) | $\geq$19 years of age | 2021-08-29 to 2021-11-27 | US | Included | mRNA | Delta | Infection & severe | Clinical | Demographic groups and those with high risk conditions for  COVID-19 disease with at least 6 events and more than 25,000 person-weeks at risk. |
| Embi (2022) | $\geq$18 years of age | 2021-01-17 to 2021-09-05 | US | Included | mRNA | Delta | Severe | Clinical | Age, geographic region, calendar time, and local virus circulation and weighted for inverse propensity to be vaccinated or unvaccinated |
| Embi2  (2021) | $\geq$18 years of age | 2021-01-17to2021-09-05 | US | Included | mRNA | Delta | Severe | Clinical | Age, geographic region, calendar time, local virus circulation, and weighted for inverse propensity to be vaccinated or unvaccinated using sociodemographic characteristics, underlying medical conditions, known previous SARS-CoV-2 infection, and hospital characteristics |
| Ferdinands (2022) | $\geq$18 years of age | 2021-08-26 to 2022-01-22 | US | Included | mRNA | Delta & omicron | Symptomatic & severe | Clinical | Age, local virus circulation, propensity to be vaccinated, and other factors |
| Fleming-Dutra  (2022) | 5-15 years of age | 2021-12-26to2022-02-21 | US | Included & excluded | mRNA | Omicron | Symptomatic | Clinical | Calendar day of test (continuous variable), race, ethnicity, sex, testing site region, and testing site census tract Social Vulnerability Index |
| Grannis (2021) | $\geq$18 years of age | 2021-06-03 to 2021-08-04 | US | Included | mRNA & Adenovirus vector vaccines | Delta | Symptomatic & severe | Clinical | Age, geographic region, calendar time, and virus circulation, and weighted for inverse propensity to be vaccinated or unvaccinated |
| Hitchings (2021) | $\geq$60 years of age | 2021-01-17 to 2021-07-02 | Brazil | Included | Adenovirus vector vaccines | Gamma | Infection & severe | Clinical | The number of reported comorbidities, previous positive SARS-CoV-2 RT-PCR or antigen test, & age |
| Husin  (2022) | 12-17 years of age | 2021-09-01to2021-12-31 | Malaysia | Included | mRNA | Delta | Infection | Non-clinical | Age, sex, states of residence, strata (urban/rural), school types, and number of baseline (before September 1, 2021) tests |
| Kim (2021) | $\geq$16 years of age | 2021-02-01 to 2021-05-28 | US | Iincluded | mRNA | Alpha | Infection | Clinical | Study site, age in years, enrollment period, race & ethnicity, and contact with a SARS-CoV-2–positive person |
| Kirsebom  (2022) | $\geq$18 years of age | 2022-01-17to2022-03-31 | UK | Included | mRNA | Omicron | Symptomatic & severe | Clinical | Age, sex, index of multiple deprivation (quintile), ethnic group, history of travel, geographic region (NHS region), period (week of test), health and social care worker status, clinical risk group status, clinically extremely vulnerable, and previously testing positive |
| Lauring (2022) | $\geq$18 years of age | 2021-03-11 to 2022-01-14 | US | Included | mRNA | Alpha & delta & omicron | Severe | Clinical | Number of  comorbidities, smoking status, living in a long term  care facility before hospital admission, and working  in a healthcare setting |
| Lewis (2021) | $\geq$18 years of age | 2021-03-11 to 2021-08-15 | US | Included | mRNA | Mix | Severe | Non-clinical | Date of admission, age, sex, self-reported race and ethnicity, burden of underlying conditions, and US Health and Human Services region of the admitting hospital |
| Lewis2  (2022) | $\geq$18 years of age | 2021-03-11to2021-12-15 | US | Included | Adenovirus vector vaccines | Alpha & delta | Severe | Non-clinical | Admission date (biweekly intervals), geographic region, age group, sex, and self-reported race and Hispanic ethnicity |
| Li (2021) | 18-59 years of age | 2021-05-18 to 2021-06-20 | China | Included | Inactivated virus vaccines | Delta | Infection & severe | Non-clinical | Age & sex |
| Maeda (2022) | $\geq$16 years of age | 2021-07-01 to 2021-09-30 | Japan | Excluded | mRNA | Delta | Infection | Clinical | Age, sex, presence of underlying medical conditions, calendar week, history of contact with COVID-19 patients, and medical institution |
| Mallow (2022) | $\geq$18 years of age | 2021-01-01 to 2021-08-25 | US | Included | mRNA | Mix | Infection | Clinical | Age, gender, race, insurance status, imputed body mass index [BMI], vaccine type, week of presentation |
| Nadeem  (2022) | $\geq$60 years of age | 2021-05-05to2021-07-31 | Pakistan | Excluded | mRNA | Delta | Infection & severe | Clinical | NA |
| Nasreen (2022) | general | 2020-12-14 to 2021-08-03 | Canada | Excluded | mRNA & Adenovirus vector vaccines | Alpha & beta & gamma & beta or gamma & delta & non-VOC | Symptomatic & severe | Clinical | Age, sex, public health unit region, period of test, number of SARS-CoV-2 tests in the 3 months before 14 December 2020, presence of any comorbidity that increase the risk of severe COVID-19, receipt of 2019/2020 and/or 2020/2021 influenza vaccination, and Census dissemination area-level quintiles of household income, proportion of persons employed as non-health essential workers, persons per dwelling, and proportion of self-identified visible minorities |
| Ngyen (2022) | general | 2020-12-23 to 2021-06-15 | France | Included | mRNA | Mix | Severe | Clinical | Time, age, and stratified on centers |
| Niessen  (2022) | $\geq$18 years of age | 2021-03-01to2021-06-26 | Netherlands | Excluded | Adenovirus vector vaccines | Alpha | Severe | Clinical | Age group and week of symptom onset |
| Olson1 (2021) | 12-18 years of age | 2021-06-01 to 2021-09-30 | US | Included | mRNA | Delta | Severe | Non-clinical | U.S. Census region, calendar month of admission, continuous age in years, sex, race or ethnicity |
| Olson2 (2022) | 12-18 years of age | 2021-07-01 to 2021-10-25 | US | Included | mRNA | Delta | Severe | Non-clinical | U.S. Census region, calendar date of admission, age, sex, and  race or ethnic group |
| Pardo-Seco (2022) | $\geq$18 years of age | 2020-12-27 to 2021-03-18 | Spain | Included | mRNA | Alpha | Infection | Non-clinical | Sex, age and time period between SARS-CoV-2 test and the start of study |
| Powell  (2022) | 12-15 years of age | 2021-09-13to2022-01-12 | UK | Excluded | mRNA | Delta & omicron | Symptomatic | Clinical | Age, sex, index of multiple deprivation (quintile), ethnic group, geographic region (NHS region), period (calendar week of onset), clinical risk group status (a separate flag for those aged over and under 16), clinically extremely vulnerable (if aged 16 and above) and previous positivity |
| Price (2022) | 12-18 years of age | 2021-07-01 to 2022-02-17 | US | Excluded | mRNA | Delta & omicron | Severe | Clinical | Sex, age, race, region, calendar time |
| Ranzani1 (2021) | $\geq$70 years of age | 2021-01-17 to 2021-04-29 | Brazil | Included | Inactivated virus vaccines | Gamma | Infection & severe | Clinical | Age and number of comorbidities |
| Ranzani2 (2022) | general | 2021-01-17 to 2021-11-27 | Brazil | Excluded | Adenovirus vector vaccines | Gamma & delta | Infection | Clinical | Age, sex, cardiovascular disease, respiratory disease, obesity, diabetes mellitus, immunosuppressed status, liver disease, occupation, region of residence, self-reported race, reason of testing, and day of the year of RT-qPCR testing symptomatic & asymptomatic |
| Rearte (2022) | $\geq$60 years of age | 2021-01-31 to 2021-09-14 | Argentina | Included | Adenovirus vector vaccines & Inactivated virus vaccines | Gamma & lambda & alpha | Infection & severe | Clinical | Epidemiological week, age, sex, history of COVID-19, and district |
| Self (2021) | $\geq$18 years of age | 2021-03-11 to 2021-08-15 | US | Included | mRNA & Adenovirus vector vaccines | Alpha & delta | Severe | Clinical | Admission date, geographic region, age, sex, and race and Hispanic ethnicity |
| Sheikh (2021) | general | 2021-04-01 to 2021-06-06 | UK | Included | mRNA & Adenovirus vector vaccines | Alpha & delta | Infection | Clinical | Age, number of prior COVID tests, date and factors for sex and deprivation |
| Skowronski  (2022) | $\geq$18 years of age | 2021-05-30to2021-11-27 | Canada | Excluded | mRNA & Adenovirus vector vaccines | Delta & alpha & gamma | Infection & severe | Non-clinical | Age group (18–49/50–69/70–79/≥80 years), sex, epi-week and region |
| Sritipsukho (2022) | $\geq$18 years of age | 2021-07-25 to 2021-10-23 | Thailand | Included | Adenovirus vector vaccines & Inactivated virus vaccines | Delta | Infection | Non-clinical | Healthcare workers, comorbidities, age, educational level, and sex |
| Suarez Castillo  (2022) | $\geq$18 years of age | 2021-12-13to2022-01-13 | France | Excluded | mRNA | Omicron/delta | Symptomatic | Clinical | Age, sex, residence, week of testing and presence of a comorbidity qualifying for prioritisation in the vaccination campaign according to the recommendations of the National Health Authority |
| Suarez  (2022) | $\geq$50 years of age | 2021-01-01to2021-12-12 | France | Excluded | mRNA | Ancestral strains & alpha & beta/gamma & delta | Symptomatic & severe | Clinical | Age (ten-year age brackets), sex, area of residence, week of testing and presence or absence of a comorbidity qualifying for prioritization in the vaccination campaign |
| Tabak (2021) | $\geq$18 years of age | 2021-05-01 to 2021-08-07 | US | Included | mRNA & Adenovirus vector vaccines | Delta | Infection | Clinical | Age, region, and calendar month of test |
| Tang (2021) | general | 2020-12-21 to 2021-09-07 | Qatar | Included | mRNA | Beta & delta | Infection & severe | Non-clinical | Prior infection & healthcare worker status |
| Tartof  (2022) | $\geq$18 years of age | 2021-12-01to2022-02-06 | US | Included | mRNA | Delta & omicron | Severe | Clinical | Age, sex, race/ethnicity, BMI, Charlson comorbidity index, prior SARS-CoV-2 infection, prior influenza vaccination, prior pneumococcal vaccination |
| Tenforde (2021) | $\geq$65 years of age | 2021-01-01 to 2021-03-26 | US | Included | mRNA | Wild type & alpha | Severe | Clinical | Region, calendar month, age, sex, and race & ethnicity |
| Tenforde2  (2022) | $\geq$18 years of age | 2021-03-11to2021-05-05 | US | Excluded | mRNA | Alpha | Severe | Clinical | Calendar time in biweekly intervals, US Department of Health and Human Services region, age in years, sex, and race and ethnicity |
| Tenforde3  (2022) | $\geq$18 years of age | 2021-03-11to2021-12-15 | US | Included | mRNA | Delta | Severe | Clinical | Calendar date of admission (in biweekly intervals), age, sex, and race and ethnicity, presence of underlying chronic conditions, immunocompromised status, and US Health and Human Services region of the admitting hospital |
| Thiruvengadam (2022) | general | 2021-04-01 to 2021-05-31 | India | Excluded | Adenovirus vector vaccines | Delta | Infection & severe | Non-clinical | Differences in age, sex, and risk of exposure to COVID-19-positive individual |
| Thompson (2021) | $\geq$50 years of age | 2021-01-01 to 2021-06-22 | US | Excluded | mRNA & Adenovirus vector vaccines | Mix | Infection & severe | Clinical | Age, geographic region, calendar time, and local virus circulation |
| Tseng (2022) | $\geq$18 years of age | 2021-12-06 to 2021-12-31 | US | Included | mRNA | Delta & omicron | Infection & severe | Non-clinical | History of SARS-CoV-2 molecular test; preventive care; number of outpatient and virtual visits; Charlson comorbidity score; obesity; frailty index; specimen type; immunocompromised status and history of COVID-19 |
| Whitaker  (2022) | $\geq$16 years of age | 2021-05-16to2021-12-07 | UK | Included | mRNA | Alpha | Symptomatic | Clinical | Week-NHS region interaction, 5-yr age group, sex, ethnicity, IMD quintile, GP record of prior COVID-19, large household, GP consultation quartile, chapter count, shielding recommendation, overall PRIMIS risk group status (overall only) and latest smoking status |
| Winkelman (2022) | $\geq$19 years of age | 2021-08-29 to 2021-10-30 | US | Included | mRNA & Adenovirus vector vaccines | Delta | Infection & severe | Non-clinical | NA |
| Zambrano (2022) | 12-18 years of age | 2021-07-01 to 2021-12-09 | US | Included | mRNA | Delta | Severe | Non-clinical | U.S. Census region, age, sex, and race & ethnicity |

**Table S3**. Summary of studies by characteristics

| Characteristics | Group | Studies |
| --- | --- | --- |
| Providing VE against infection | NA | (1-51) |
| Providing VE against severe disease | NA | (1, 2, 6, 10, 12-14, 18-21, 26, 28, 30, 34-38, 40, 44, 45, 47, 49, 50, 52-67) |
| COVID-19 infection history | Included | (1, 2, 7-10, 13, 14, 16, 18-23, 25, 27, 28, 30-34, 37, 38, 40-44, 50-58, 60-64, 67) |
|  | Excluded | (3-6, 10-12, 15, 17, 24, 26, 29, 35, 36, 39, 42, 45-49, 59, 65, 66) |
| Enrolment criteria | Clinical | (3, 5-9, 12, 15, 16, 18-22, 24-26, 28-31, 33, 36, 40, 42, 44-46, 48-51, 53, 54, 56, 59-61, 63, 65-67) |
|  | Non-clinical | (1, 2, 4, 10, 11, 13, 14, 17, 23, 27, 32, 34, 35, 37-39, 41, 43, 47, 52, 55, 57, 58, 62, 64) |
| Vaccine type for VE against infection | mRNA vaccines | (1-11, 13-20, 22, 24-27, 31, 33, 34, 36-63, 65-67) |
|  | Adenovirus vector vaccines | (3, 5-7, 9, 20, 21, 26, 29-33, 35, 36, 38, 40, 47, 60, 64) |
|  | inactivated virus vaccines | (12, 23, 28, 30, 32) |
| Vaccine type for VE against severe disease | mRNA vaccines | (1, 2, 6, 10, 13, 14, 18-20, 26, 34, 36-38, 40, 44, 45, 47, 49, 50, 52-63, 65-67) |
|  | Adenovirus vector vaccines | (6, 20, 21, 26, 30, 35, 36, 38, 40, 47, 60, 64) |
|  | inactivated virus vaccines | (12, 28, 30) |
| Circulating virus type for VE against infection | Omicron only | (5, 37, 39, 41, 42, 44, 46, 48, 50, 52, 54, 59) |
|  | Delta only and Delta with Omicron | (5-7, 9, 10, 17-20, 23, 24, 26, 31, 32, 34, 35, 37, 38, 40, 43, 45-50, 52-54, 57-59, 62, 63, 67) |
|  | Delta with non-omicron variants and ancestral strains | (1, 12, 13, 25, 29, 33, 55, 60, 64) |
|  | Ancestral strains and variants except Delta and Omicron | (2-4, 6-11, 14-17, 21, 22, 26-28, 30, 31, 34, 36, 47, 49, 51, 54, 56, 61, 65, 66) |
| Circulating virus type for VE against severe disease | Omicron only | (37, 44, 50, 52, 54, 59) |
|  | Delta only and Delta with Omicron | (2, 10, 18-20, 26, 34, 35, 37, 38, 40, 45, 47, 49, 50, 52-54, 57-59, 62, 63, 67) |
|  | Delta with non-omicron variants and ancestral strains | (1, 12, 13, 55, 60, 64) |
|  | Ancestral strains and variants except Delta and Omicron | (2, 6, 14, 21, 26, 28, 30, 34, 36, 47, 49, 54, 56, 61, 65, 66) |

**Table S4.** Summary of number of studies and estimates by factors

|  | Infection | | Severe disease | |
| --- | --- | --- | --- | --- |
|  | Number of studies | Number of estimates | Number of studies | Number of estimates |
| Handling of participants with prior infection | | | | |
| Included | 32 | 77 | 30 | 53 |
| Excluded | 21 | 96 | 11 | 40 |
| Enrolment criteria | | | | |
| Clinical | 25 | 115 | 25 | 59 |
| Non-clinical | 17 | 58 | 17 | 34 |
| Vaccine type |  |  |  |  |
| mRNA | 42 | 126 | 34 | 73 |
| Adenovirus vector | 19 | 41 | 12 | 16 |
| Inactivated | 6 | 6 | 4 | 4 |
| Predominant circulating virus | | | | |
| Pre-Delta | 25 | 92 | 16 | 35 |
| Delta | 6 | 9 | 6 | 8 |
| Late-Delta | 26 | 54 | 24 | 42 |
| Omicron | 10 | 18 | 7 | 8 |

**Table S5.** Number of estimates available disaggregated by handling of participants with prior infection

|  | Included participants with prior infection | | | | Excluded participants with prior infection | | | |
| --- | --- | --- | --- | --- | --- | --- | --- | --- |
|  | Infection | | Severe disease | | Infection | | Severe disease | |
|  | Number of studies | Number of estimates | Number of studies | Number of estimates | Number of studies | Number of estimates | Number of studies | Number of estimates |
| Handling of participants with prior infection |  |  |  |  |  |  |  |  |
| Included | 32 | 77 | 30 | 53 |  |  |  |  |
| Excluded |  |  |  |  | 21 | 96 | 11 | 40 |
| Enrolment criteria |  |  |  |  |  |  |  |  |
| Clinical | 19 | 49 | 16 | 32 | 14 | 66 | 9 | 27 |
| Non-clinical | 13 | 28 | 14 | 21 | 7 | 30 | 3 | 13 |
| Vaccine type |  |  |  |  |  |  |  |  |
| mRNA | 27 | 56 | 26 | 43 | 17 | 70 | 8 | 30 |
| Adenovirus vector | 11 | 17 | 7 | 8 | 8 | 24 | 5 | 8 |
| Inactivated | 4 | 4 | 2 | 2 | 2 | 2 | 2 | 2 |
| Predominant circulating virus |  |  |  |  |  |  |  |  |
| Pre-Delta | 15 | 32 | 9 | 12 | 11 | 60 | 7 | 23 |
| Delta | 4 | 7 | 5 | 7 | 2 | 2 | 1 | 1 |
| Late-Delta | 15 | 29 | 17 | 27 | 12 | 25 | 7 | 15 |
| Omicron | 6 | 9 | 6 | 7 | 5 | 9 | 1 | 1 |

**Table S6.** Relationship between pre-existing population immunity and estimates of risk ratios against infection or death, using different measures.

| Endpoint | Infection | Severe disease |
| --- | --- | --- |
| *Model 1: adjusted for vaccine type, circulating virus and recruitment criteria + age group (baseline factors) + study including participants with COVID infection history* | | |
| Vaccine type |  |  |
| mRNA vaccines | ref | ref |
| Adenovirus vector vaccines | 2.24 (1.79, 2.8) | 1.71 (1.12, 2.6) |
| inactivated virus vaccines | 2.07 (1.26, 3.39) | 3.1 (1.5, 6.41) |
| Predominant circulating virus |  |  |
| pre-Delta | ref | ref |
| Delta | 2.16 (1.36, 3.41) | 1.36 (0.71, 2.62) |
| late Delta | 1.85 (1.5, 2.3) | 1.12 (0.77, 1.64) |
| Omicron | 5.95 (4.26, 8.31) | 3.81 (2.09, 6.96) |
| Enrolment criteria |  |  |
| Non-clinical | ref | ref |
| Clinical | 1.19 (0.96, 1.47) | 1.59 (1.15, 2.21) |
| Including participants with COVID infection history | 1.56 (1.29, 1.89) | 1.73 (1.23, 2.45) |
| *Model 2: Baseline factors + cumulative incidence of cases before start of study* | | |
| Doubling cumulative incidence of cases | 1.1 (1.02, 1.2) | 1.47 (1.26, 1.71) |
| *Model 3: Baseline factors + incidence of cases during study* | | |
| Doubling incidence of cases | 1.16 (1.07, 1.25) | 1.2 (1.03, 1.4) |

**Table S7.** Sensitivity analysis for relationship between pre-existing population immunity and estimates of risk ratios against infection or death, using different measures.

| Endpoint | Infection | Severe disease |
| --- | --- | --- |
| *Model 1: adjusted for vaccine type, circulating virus and recruitment criteria + duration of study + age group + countries (US or US or others) (baseline factors) + study including participants with COVID infection history* | | |
| Vaccine type |  |  |
| mRNA vaccines | ref | ref |
| Adenovirus vector vaccines | 2.18 (1.74, 2.75) | 1.98 (1.38, 2.83) |
| inactivated virus vaccines | 1.88 (1.12, 3.15) | 3.74 (1.96, 7.12) |
| Predominant circulating virus |  |  |
| pre-Delta | ref | ref |
| Delta | 2.09 (1.3, 3.36) | 1.26 (0.7, 2.26) |
| late Delta | 1.87 (1.49, 2.34) | 1.08 (0.76, 1.52) |
| Omicron | 6.16 (4.24, 8.94) | 3.5 (1.96, 6.27) |
| Enrolment criteria |  |  |
| Non-clinical | ref | ref |
| Clinical | 1.23 (0.96, 1.57) | 1.42 (1.01, 1.98) |
| Including participants with COVID infection history | 1.71 (1.39, 2.12) | 1.34 (0.98, 1.84) |
| *Model 2: Baseline factors + cumulative incidence of cases before start of study* | | |
| Doubling cumulative incidence of cases | 1.15 (1.05, 1.26) | 1.25 (1, 1.55) |
| *Model 3: Baseline factors + incidence of cases during study* | | |
| Doubling incidence of cases | 1.17 (1.07, 1.27) | 1.12 (0.96, 1.31) |

**Table S8.** Meta-analysis disaggregated by handling of participants with prior infection

|  | Infection | | Severe disease | |
| --- | --- | --- | --- | --- |
|  | Excluding prior infection | Including prior infection | Excluding prior infection | Including prior infection |
| Vaccine type | | | | |
| mRNA vaccines | 90 (87, 91) | 81 (75, 85) | 94 (92, 96) | 88 (85, 91) |
| Adenovirus vector vaccines | 74 (69, 79) | 62 (55, 69) | 91 (85, 95) | 78 (74, 82) |
| Inactivated virus vaccines | 84 (-22, 98) | 49 (40, 57) | 76 (50, 89) | 66 (44, 79) |
| Predominant circulating virus | | | | |
| pre-Delta | 90 (88, 92) | 86 (80, 89) | 93 (90, 95) | 91 (80, 96) |
| Delta | 55 (54, 56) | 81 (60, 91) | 82 (81, 83) | 88 (75, 94) |
| late Delta | 86 (83, 89) | 70 (63, 75) | 95 (92, 97) | 87 (83, 90) |
| Omicron | 59 (48, 68) | 44 (35, 52) | 20 (-25, 49) | 66 (58, 72) |
| Enrolment criteria | | | | |
| Clinical | 85 (82, 87) | 75 (69, 80) | 92 (88, 94) | 82 (78, 86) |
| Non-clinical | 91 (87, 94) | 80 (71, 87) | 95 (93, 97) | 92 (87, 95) |

47.	Skowronski DM, Febriani Y, Ouakki M, Setayeshgar S, El Adam S, Zou M, et al. Two-Dose Severe Acute Respiratory Syndrome Coronavirus 2 (SARS-CoV-2) Vaccine Effectiveness With Mixed Schedules and Extended Dosing Intervals: Test-Negative Design Studies From British Columbia and Quebec, Canada. Clinical Infectious Diseases.
